# Public Views about COVID-19 “Immunity Passports”

**DOI:** 10.1101/2021.01.26.21250184

**Authors:** Mark A. Hall, David M. Studdert

## Abstract

**Importance:** Discovery of effective vaccines and increased confidence that infection confers extended protection against COVID-19 have renewed discussion of using immunity certificates or “passports” to selectively reduce ongoing public health restrictions.

**Objective:** To determine public views regarding government and private conferral of immunity privileges.

**Design and Setting:** National on-line survey fielded in June 2020. Participants were randomly asked about either government “passports” or private “certificates” for COVID-19 immunity.

**Participants:** Adults from a standing panel maintained for academic research, selected to approximate national demographics.

**Main Outcomes/Measures:** Level of support/opposition to immunity privileges, and whether views vary based on: government vs. private adoption; demographics; political affiliation or views; or various COVID19-related attitudes and experiences.

**Results:** Of 1315 respondents, 45.2% supported immunity privileges, with slightly more favoring private certificates than government passports (48.1% vs 42.6%, p=0.04). Support was greater for using passports or certificates to enable returns to high-risk jobs or attendance at large recreational events than for returning to work generally. Levels of support did not vary significantly according to age groups, socioeconomic or employment status, urbanicity, political affiliation or views, or whether the respondent had chronic disease(s). However, estimates from adjusted analyses showed less support among women (Odds Ratio, 0.64; 95% Confidence Interval, 0.51 to 0.80), and among Hispanics (0.56; 0.40 to 0.78) and other minorities (0.58; 0.40 to 0.85) compared with whites, but not among blacks (0.83; 0.60 to 1.15). Support was much higher among those who personally wanted a passport or certificate (75.6% vs 24.4%) and much lower among those who believed this would harm the social fabric of their community (22.9% vs 77.1%).

**Conclusions and Relevance:** Public views are divided on either government or private use of immunity certificates, but, prior to any efforts to politicize the issues, these views do not vary along usual political lines, nor by characteristics that indicate individual vulnerability to infection. Social consensus on the desirability of an immunity privileges programs may be difficult to achieve.

**Key Points:** *Question:* What are the public’s views on government or private use of immunity “passports” to selectively lift COVID-19 restrictions?

*Findings:* Views are divided and do not vary substantially according to political affiliation or many demographic factors. Support is greater among men but lower among Hispanics and those who believe that immunity privileges would harm the social fabric of society.

*Meaning:* Social consensus will be difficult to achieve on the appropriateness of immunity privileges.

---

Earlier in the COVID-19 pandemic, the idea of immunity certifications (or “passports”) was introduced as a means to lift public health restrictions on recovered patients who might be considered safe from reinfection or further viral spread.^1,2^ Despite some thoughtful support,^3,4^ the idea quickly encountered substantial opposition, based partly on concerns about social fairness.^5,6^ Scientific uncertainty over the extent of acquired immunity was another chilling factor. However, there have been extraordinarily few documented reinfections to date^7,8^ and more recent studies indicate that even mild infection confers some sustained viral defense.^9,10^

This evidence, coupled with new rounds of restrictions as COVID-19 cases surge, is likely to prompt renewed calls for easing restrictions on those who can establish likely immunity.

Tailoring restrictions according to individual risk could help to defend public health restrictions from legal or public opposition.^11^ Also, as vaccines are rolled out, easing restrictions on the vaccinated could encourage vaccination and speed returns to normalcy.

Even without official exemptions, there are signs that immunity certification may advance through private initiative.^12^ Firms are developing smart technologies that allow recovered or vaccinated individuals to verify their presumptively safe status.^4^ Airlines are considering immunity certification to promote safer travel. And employers face economic and regulatory pressures to triage higher-exposure functions.^1^ We sought to gauge public views about government or private use of immunity certification.

## Methods

We conducted a national survey in late June 2020 using the on-line survey panel, Prolific Academic, which has demonstrated good reliability and validity in prior studies.^4,13,14^ We selected 1315 respondents using a quota system that approximated nationally-representative demographics by sex, age groups, and race-ethnicity (eTable 1).

The survey randomly split respondents into two arms -- one that described government adoption of an immunity “passport” and the other private adoption of an immunity “certificate.” Each arm explained the immunity privilege concept as follows: “if an antibody test shows that you have had the disease, you could receive an ‘[immunity passport/certificate]’ which would let you engage in more activities.”

Support for the concept was assessed following questions about the fairness, acceptability, and potential drawbacks of using passports/certificates for various specified purposes. Respondents also reported experiences with COVID-19 and related restrictions, along with residential location, employment, socio-economic status, health, and political affiliations. The full questionnaire appears in the Appendix.

We calculated counts and proportions to describe support for immunity privileges, by demographic subgroups and by attitudinal and experiential variables. Respondents indicated their degree of support or opposition on a 6-point Likert scale; we dichotomized this variable, classifying “Strongly support”, “Support” and “Somewhat support” responses as support.

Finally, we used multivariable logistic regression to estimate associations between support and demographic characteristics.

Statistical analyses were performed using Stata, version 14.1.

## Results

Respondents were fairly evenly split in their support for immunity privileges (Figure 1). Nearly half (45.2%) supported them, with more in favor of private certificates than government passports (48.1% vs 42.6%, p=0.04). Respondents were more likely to view passports or certificates as fair for determining who may return to high-risk jobs or attend large recreational events than for returning to work generally (Figure 1).

**Figure 1.**
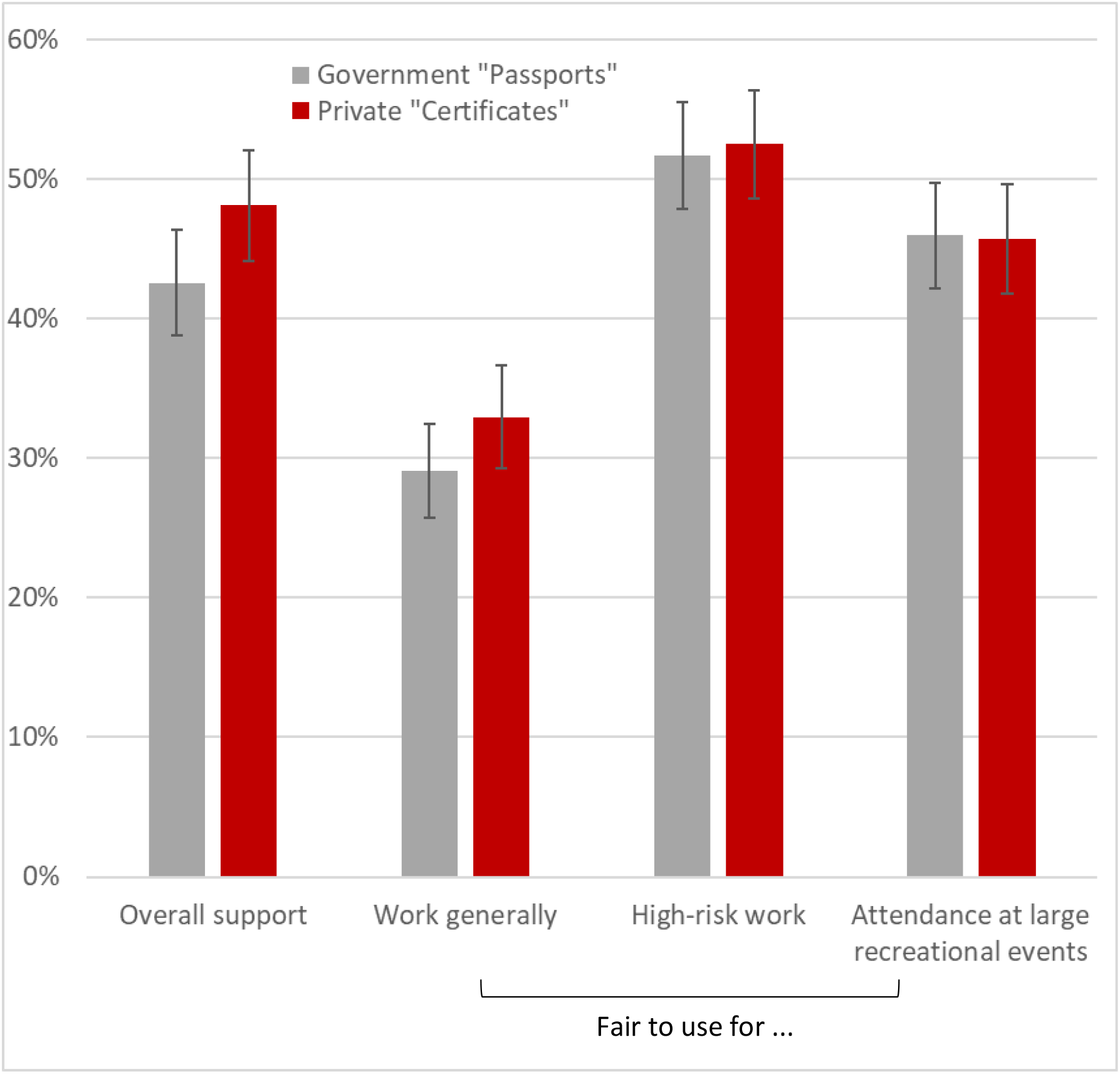
Support for immunity privileges and perceived fairness of their use by activity.

The overall level of support for immunity privileges was similar across most demographic groups examined (Table 1). Estimates from the adjusted analysis indicated less support among women (Odds Ratio, 0.64; 95% Confidence Interval, 0.51 to 0.80), and among Hispanics (0.56; 0.40 to 0.78) and other minorities (0.58; 0.40 to 0.85) compared with whites, but not among blacks (0.83; 0.60 to 1.15). Support did not vary significantly according to political affiliation or characteristics that mark vulnerability to COVID-19, such as age, chronic disease, low socioeconomic status, and customer-facing employment.

**Table 1.**
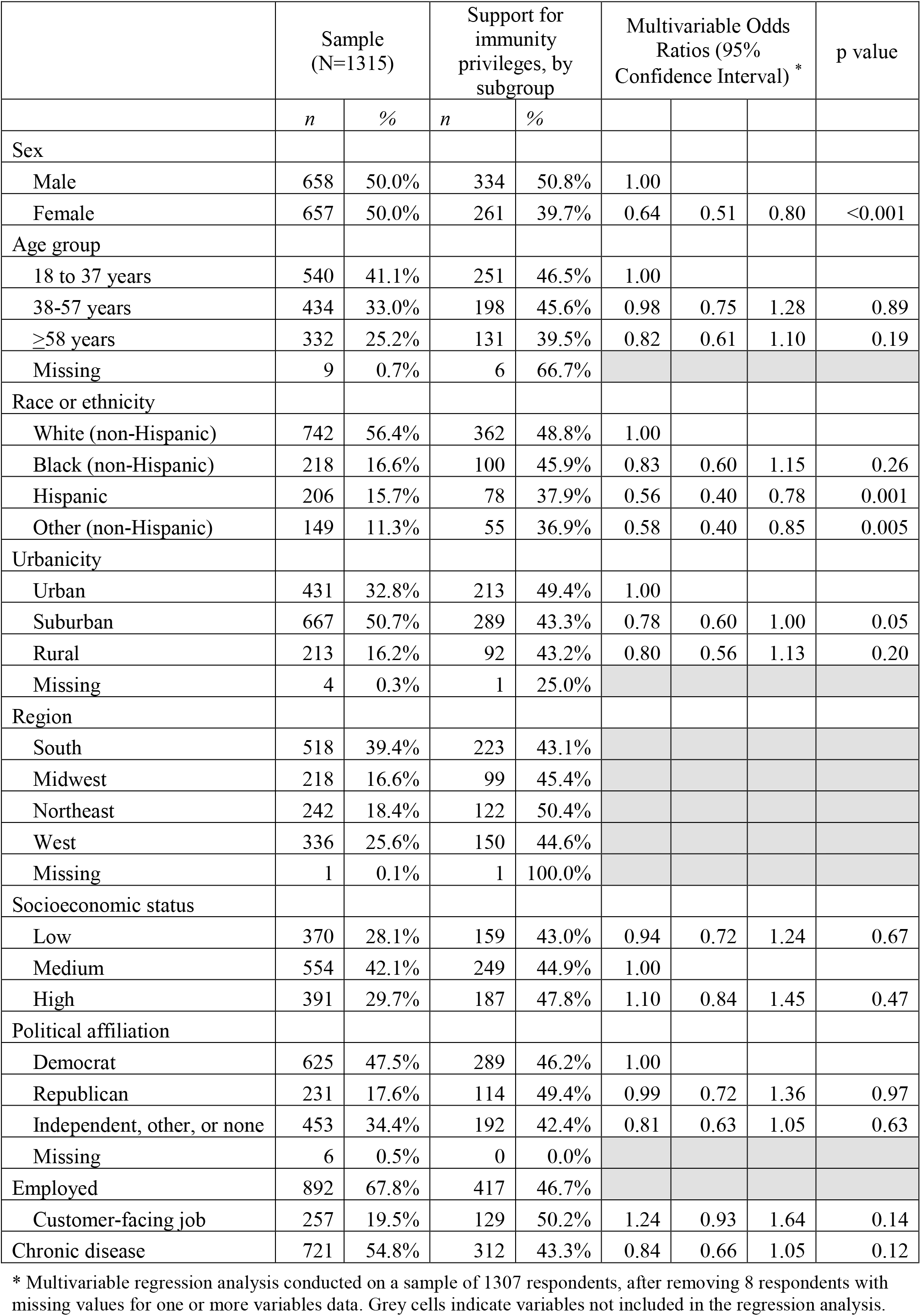
Support for immunity privileges, by demographic subgroups.

|  | Sample<br>(N=1315) |  | Support for<br>immunity<br>privileges, by<br>subgroup |  | Multivariable Odds<br>Ratios (95%<br>Confidence Interval) * |  |  | p value |
| --- | --- | --- | --- | --- | --- | --- | --- | --- |
|  | *n* | % | *n* | % |  |  |  |  |
| Sex |  |  |  |  |  |  |  |  |
| Male | 658 | 50.0% | 334 | 50.8% | 1.00 |  |  |  |
| Female | 657 | 50.0% | 261 | 39.7% | 0.64 | 0.51 | 0.80 | <0.001 |
| Age group |  |  |  |  |  |  |  |  |
| 18 to 37 years | 540 | 41.1% | 251 | 46.5% | 1.00 |  |  |  |
| 38-57 years | 434 | 33.0% | 198 | 45.6% | 0.98 | 0.75 | 1.28 | 0.89 |
| ≥58 years | 332 | 25.2% | 131 | 39.5% | 0.82 | 0.61 | 1.10 | 0.19 |
| Missing | 9 | 0.7% | 6 | 66.7% |  |  |  |  |
| Race or ethnicity |  |  |  |  |  |  |  |  |
| White (non-Hispanic) | 742 | 56.4% | 362 | 48.8% | 1.00 |  |  |  |
| Black (non-Hispanic) | 218 | 16.6% | 100 | 45.9% | 0.83 | 0.60 | 1.15 | 0.26 |
| Hispanic | 206 | 15.7% | 78 | 37.9% | 0.56 | 0.40 | 0.78 | 0.001 |
| Other (non-Hispanic) | 149 | 11.3% | 55 | 36.9% | 0.58 | 0.40 | 0.85 | 0.005 |
| Urbanicity |  |  |  |  |  |  |  |  |
| Urban | 431 | 32.8% | 213 | 49.4% | 1.00 |  |  |  |
| Suburban | 667 | 50.7% | 289 | 43.3% | 0.78 | 0.60 | 1.00 | 0.05 |
| Rural | 213 | 16.2% | 92 | 43.2% | 0.80 | 0.56 | 1.13 | 0.20 |
| Missing | 4 | 0.3% | 1 | 25.0% |  |  |  |  |
| Region |  |  |  |  |  |  |  |  |
| South | 518 | 39.4% | 223 | 43.1% |  |  |  |  |
| Midwest | 218 | 16.6% | 99 | 45.4% |  |  |  |  |
| Northeast | 242 | 18.4% | 122 | 50.4% |  |  |  |  |
| West | 336 | 25.6% | 150 | 44.6% |  |  |  |  |
| Missing | 1 | 0.1% | 1 | 100.0% |  |  |  |  |
| Socioeconomic status |  |  |  |  |  |  |  |  |
| Low | 370 | 28.1% | 159 | 43.0% | 0.94 | 0.72 | 1.24 | 0.67 |
| Medium | 554 | 42.1% | 249 | 44.9% | 1.00 |  |  |  |
| High | 391 | 29.7% | 187 | 47.8% | 1.10 | 0.84 | 1.45 | 0.47 |
| Political affiliation |  |  |  |  |  |  |  |  |
| Democrat | 625 | 47.5% | 289 | 46.2% | 1.00 |  |  |  |
| Republican | 231 | 17.6% | 114 | 49.4% | 0.99 | 0.72 | 1.36 | 0.97 |
| Independent, other, or none | 453 | 34.4% | 192 | 42.4% | 0.81 | 0.63 | 1.05 | 0.63 |
| Missing | 6 | 0.5% | 0 | 0.0% |  |  |  |  |
| Employed | 892 | 67.8% | 417 | 46.7% |  |  |  |  |
| Customer-facing job | 257 | 19.5% | 129 | 50.2% | 1.24 | 0.93 | 1.64 | 0.14 |
| Chronic disease | 721 | 54.8% | 312 | 43.3% | 0.84 | 0.66 | 1.05 | 0.12 |

\* Multivariable regression analysis conducted on a sample of 1307 respondents, after removing 8 respondents with missing values for one or more variables data. Grey cells indicate variables not included in the regression analysis.

However, supporters and opposers differed in several, but not all, of the attitudinal and experiential factors shown in Figure 2. For example, respondents who reported that they wanted a passport or certificate were substantially more likely to support the concept (75.6% vs 24.4%). On the other hand, those who believed these programs would harm the social fabric of their community (22.9% vs 77.1%) or that it may be years before we have a “safe and effective vaccine generally available” (37.7% vs 62.3%) were substantially more likely to oppose.

**Figure 2.**
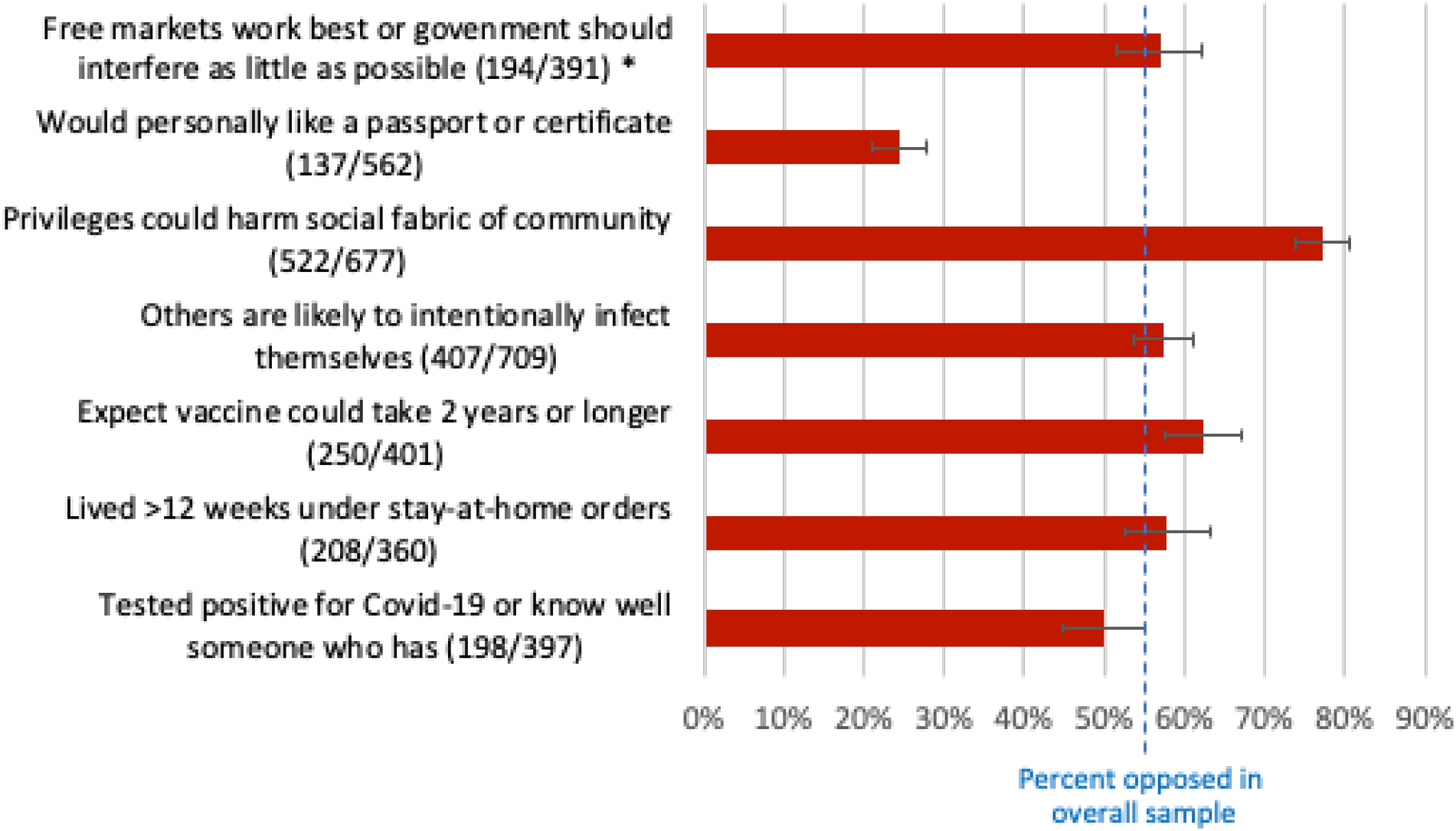
Percent opposed to immunity privileges, by other views and experiences ^*^ Number who oppose immunity privileges among total respondents with each view or experience.

## Discussion

This nationally-representative survey, conducted in the early Summer of 2020, as the first round of COVID-19 restrictions was lifting, found the public was more-or-less evenly divided on the appropriateness of using immunity privileges programs to selectively allow people to return to normal, pre-pandemic activities. Overall, 55% of respondents opposed the idea, although fewer opposed certifications authorized by the private sector than by government. Levels of support did not vary across many of the characteristics that frequently mark divergent views about social and public health policies.

In mid-2020, political affiliation was not associated with support for or opposition to immunity privileges. This finding is particularly noteworthy considering how deeply politicized so many aspects of COVID-19 public health policy have been. One explanation is that the “immunity passport” idea has not yet become sufficiently aired for political camps to form a position that could influence the broader public. Another explanation is that, prior to any efforts to politicize the issue, immunity programs were seem as having pros and cons that cross political lines: conservatives may welcome the potential boost to economic recovery while lamenting identity-specific control of civil liberties; progressives may resent giving selective privileges, but may also recognize the potential for those who have fared worse—minorities and the poor—to gain the most.

Men were significantly more likely to support immunity privileges, as were non-Hispanic whites and blacks. But systematic differences were not evident across other demographic groupings we examined. Nor did we find that vulnerability to COVID-19—based on older age or lower health or socioeconomic status—explained support for immunity privileges.

On the other hand, certain attitudes were strongly associated with opposition to immunity privileges. For example, those who thought that immunity privileges would harm the community’s social fabric were more likely to oppose them. Somewhat counterintuitively, opposers were also more likely among those who, at the time of this survey, expected a longer wait for a safe and effective vaccine. That alignment of views could reflect a desire for greater social solidarity.

Our results are limited by the standard validity and reliability concerns that attend on-line attitudinal surveys, although the panel we used was developed for academic research. We fielded this cross-sectional survey in mid-2020, when the pandemic was at a different stage, and respondents’ beliefs may have changed since then. Among the potentially influential changes since the original survey, which could affect current views, is the increasing widespread availability of immunity via a safe and effective vaccine, which additionally makes it much less likely that people might self-infect in order to acquire immunity.

Also, respondents’ views may have been sensitive to assumptions about the accuracy of immunity testing or completeness of immunity protection. We probed this latter issue through a supplemental randomized experiment and found significantly higher support following an assurance that immunity protection is virtually (99%) certain, compared with no information on this point, but not when the assurance indicated lower levels of certainty (80%, 90%) (eTable 2).

Today more than 50 million Americans may be immune to SARS-CoV-2 from prior infection.^15^ The roll-out of vaccines in 2021 will gradually confer strong protection to many millions more. Meanwhile, the Winter spike in COVID-19 cases is forcing shut-downs across the country. This confluence will inevitably reignite debate about selectively relaxing restrictions. The public appears divided over whether doing so is appropriate, although, in mid-2020, prior to efforts to politicize the issue, the division did not appear to be along typical political and social lines.

## Data Availability

Data is available by contacting the corresponding author

