## Supplemental Appendix for "Public Views about COVID-19 “Immunity Passports”" (from another version of this preprint: version 1, 2021-01-27)

**Supplementary Appendix**

| **eTable 1. Demographic Characteristics of Study Sample Compared with National Averages** | |  |
| --- | --- | --- |
|  | **Study Sample** | **National** |
| Sex |  |  |
| Male | 51% | 49% |
| Female | 49% | 51% |
| Age group |  |  |
| 18 to 37 years | 46% | 35% |
| 38-57 years | 31% | 33% |
| ≥58 years | 21% | 32% |
| Race or ethnicity |  |  |
| White (nonHispanic) | 58% | 63% |
| Black (nonHispanic) | 15% | 12% |
| Hispanic | 15% | 16% |
| Other (nonHispanic) | 12% | 9% |
| Urbanicity |  |  |
| Urban | 33% | 31% |
| Surburban | 50% | 55% |
| Rural | 16% | 14% |
| Region |  |  |
| South | 40% | 38% |
| Northeast | 18% | 17% |
| West | 25% | 24% |
| Midwest | 17% | 21% |
| Political affiliation |  |  |
| Democrat | 45% | 33% |
| Republican | 22% | 26% |
| Independent, other, or none | 32% | 41% |

**Supplemental Survey Testing Effects of Stating Specific Accuracy Levels for Immunity Testing**

We conducted a supplemental survey designed to test experimentally whether the respondents’ answers to the primary survey were affected by their concerns about either the accuracy of antibody testing or the degree of protection conferred by coronavirus antibodies.

Using the same survey panel (Prolific Academic), we recruited 1420 subjects who were not respondents to the primary survey. These subjects were randomly assigned to one of four experimental arms. The first arm essentially repeated the initial survey, including its description of immunity privileges, only leaving out any distinction between government passports versus private certificates by describing them simply as “immunity certificates” without specifying their source. The other three arms described these as “time limited” certificates and added this additional explanation:

Currently, experts are unsure how effective immunity is to COVID-19 coronavirus. However, for purposes of this survey please ASSUME that antibody testing will be up to ___ percent accurate in detecting true immunity. That means if someone tests positive for antibodies, assume there is a ___% chance they can**not** get sick with COVID-19 **or** spread it to others, but a ___% chance it's possible they could. Also, assume this protection will last at least as long as the certificate covers.

The stated percentages were randomly varied to be either 80/20, 90/10, or 99/1.

Because many of the panel’s older and minority members had been sampled in the primary survey, the demographic composition of respondents to this supplemental survey was not nationally representative. That mismatch is of greater concern in a study aiming to reflect the level of support nationally, but that was not the aim of this supplemental survey. Instead, we sought to determine through a pure experimental design whether altered wording has the potential to change views. Other studies have shown that convenience samples such as this are well suited to that purpose.^1–3^

eTable 2 reports key results from the supplemental survey. Support for immunity certificates ranged from 32.3% among respondents in the arm in which the certainty of immunity protection was not specified, to 43.9% among respondents in the arm where 99% protection was specified. (Note that the level of support in the “unstated” arm was lower than that found among respondents to the primary survey, which may reflect differences in the demographic composition of the two study samples.) A chi-square test for differences across these groups was significant (P=0.02). Additional significance testing across selected strata suggested this difference was driven largely by the relatively high level of support among respondents in the “99% effective” arm.

**eTable 2. Support for immunity certificates according to degree of certainty of immunity protection**

| **Certainty of immunity protection** | **Support Immunity Certificates** ^*^ | | **Odds Ratio** ^†^  **(95% Confidence Interval)** | |
| --- | --- | --- | --- | --- |
|  | ***n*** | ***%*** |  |  |
| Unstated *(n=356)* | 115 | 32.3% | 1.00 | *Reference* |
| 80% certain *(n=356)* | 132 | 37.1% | 1.22 | (0.90 to 1.67) |
| 90% certain *(n=353)* | 130 | 36.8% | 1.24 | (0.90 to 1.69) |
| 99% certain *(n=355)* | 156 | 43.9% | 1.64 | (1.20 to 2.23) |
| ^*^ Respondents indicated their degree of support or opposition on a 6-point Likert scale; we dichotomized this variable, classifying “Strongly support”, “Support” and “Somewhat support” responses as support.  ^†^ Analysis adjusts for respondents’ sex, age group, race or ethnicity, SES, political party affiliation, employment in a customer-facing job, and chronic health condition. Specification of these variables followed that used in the multivariable analyses reported in Table 1 of the manuscript. | | | | |

eTable 2 also reports results of a logistic regression analysis of the extent to which the degree/certainty of immunity is associated with support for immunity certificates. The dependent variable in this analysis is specified in the same way as the “support” variable discussed above (see note beneath table) and the analysis adjusted for the same covariates that were included in the multivariable regression reported in Table 1 of the manuscript. Respondents in the 99% arm had higher odds of supporting immunity certificates than respondents in the “unstated” arm. Respondents in the 80% and 90% arms also had odds ratios greater than 1, but the confidence intervals crossed 1.

In sum, we detected evidence that stating that “immunity” confers near-certain protection produces higher levels of support for immunity certificates, compared with offering no indication of the degree of protection. However, stating high but lesser levels of protection (80% or 90%) did not significantly affect support.

**Survey Questions**

1. Have you ever been tested for COVID-19 coronavirus or its antibodies?
2. [If yes]: Have you received a positive test result, indicating that you have or have had COVID-19 (coronavirus) or its antibodies?
3. Has somebody you know well ever tested positive for COVID-19 coronavirus or its antibodies? [Yes, No]
4. How many weeks, if any, have you had to stay at home due to restrictions related to the COVID-19 coronavirus? ___ weeks [fill in whole number]
5. Using your best guess, how long do you think it will be before we have a safe and effective vaccine generally available for the COVID-19 coronavirus? [about 6 months, closer to a year, about 18 months, up to 2 years, more than 2 years, Never.]

[Respondents were randomly assigned to one of two arms -- one (“p”) that described government adoption of an immunity “passport” and the other (“c”) private adoption of an immunity “certificate.” Each arm explained the concept as follows:]

Immunity [passports/certificates] have been proposed as a potential step towards the [government/private businesses] lifting restrictions during the COVID-19 pandemic, especially if the pandemic worsens again. An '[immunity passport/certificate]’ indicates that you have had a disease and that you have the antibodies for the virus causing that disease. Having the antibodies to COVID-19 (coronavirus) implies that you are now immune and therefore not able to spread the virus to other people. Thus, if an antibody test shows that you have had the disease, you could receive an '[immunity passport/certificate]' which would let you engage in more activities.

6. How much would you like to have an [immunity passport/certificate] for COVID-19 coronavirus? [Not at all to Extremely – 6 points]
7. To what extent do you believe an [immunity passport/certificate] for COVID-19 coronavirus could harm the social fabric of your community?  [Not at all to Extremely – 6 points]
8. How fair or unfair would it be for [government/private employers] to allow people to go to work only if they have an [immunity passport/certificate] for COVID-19 coronavirus?  [Extremely unfair to extremely fair – 6 points]
9. How fair or unfair would it be for [government/private employers] to allow people to work in high-risk jobs, such as personal care or meat packing, only if they have an [immunity passport/certificate] for COVID-19 coronavirus? [Extremely unfair to extremely fair – 6 points]
10. How fair or unfair would it be for [government/private businesses] to allow people to attend large recreational events [they host] like concerts or sports games only if they have an [immunity passport/certificate] for COVID-19 coronavirus?  [Extremely unfair to extremely fair – 6 points]
11. How likely would you be to intentionally infect yourself with the COVID-19 coronavirus to get an [immunity passport/certificate] for COVID-19 coronavirus?  [Extremely unlikely to Extremely Likely– 6 points]
12. How likely do you think a lot of other people would intentionally infect themselves with the COVID-19 coronavirus to get an [immunity passport/certificate]?  [Extremely unlikely to Extremely Likely – 6 points]
13. Would you support or oppose [a government proposal to introduce [immunity passport/private businesses recognizing immunity certificates] for COVID-19 coronavirus?  [6 point Strongly Oppose to Strongly Support, with no undecided,]
14. Free markets that are unrestrained by government work best to meet human needs. [7-point Strongly Disagree to Strongly Agree]
15. The government should interfere with the lives of citizens as little as possible. [7-point Strongly Disagree to Strongly Agree]

END OF SURVEY. THE REMAINING ITEMS WERE PROVIDED BY THE SURVEY FIRM, BASED ON INFORMATION PREVIOUSLY COLLECTED FROM RESPONDENTS

22. In general, what is your political affiliation? [Democrat, Republican, Independent, Other, None]
23. Age
24. Sex
30. Race/Ethnicity [White/Caucasian; Black/African-American; Hispanic/Latino; Other]
31. In what state do you live?
32. How would you describe the community where you live? [1=Urban, 2=Suburban 3=Rural or small town]
33. Think of a ladder (see image) as representing where people stand in society. At the top of the ladder are the people who are best off—those who have the most money, most education and the best jobs. At the bottom are the people who are worst off—who have the least money, least education and the worst jobs or no job. The higher up you are on this ladder, the closer you are to people at the very top and the lower you are, the closer you are to the bottom. Where would you put yourself on the ladder? Choose the number [from 10-1] whose position best represents where you would be on this ladder.
35. Do you have – or have you had – any condition, injury, or chronic illness?

- Autoimmune diseases
- Cancers
- Diabetes
- Genetics/Birth Defects
- Infections
- Injuries and Wounds
- Mental Health and Behavior
- Metabolic Problems
- Poisoning, Toxicology, Environmental Health
- Pregnancy and Reproduction
- Substance Abuse Problems
- Other
- None/Rather not say

36. Are you employed in a customer facing\ front line employee role such as retail or banking? [Yes, No, Don't know/ Rather not say]
37. What is your employment status

- Full-Time
- Part-Time
- Due to start a new job within the next month
- Unemployed (and job seeking)
- Not in paid work (e.g. homemaker, retired or disabled)
- Other
